# Differentiating radiation necrosis from recurrent brain metastases using magnetic resonance elastography

**DOI:** 10.64898/2026.03.04.26347674

**Authors:** Jan Saip Aunan-Diop, Ancuta Ioana Friismose, Ziying Yin, Emi Hojo, Jeanette Krogh Pettersen, Mads Hjortdal Grønhøj, Christian Bonde Pedersen, Bo Mussmann, Bo Halle, Frantz Rom Poulsen

## Abstract

**Background:** Conventional MRI cannot reliably distinguish radiation necrosis (RN) from recurrent metastasis after cranial radiotherapy, as both can show similar enhancement despite different biology. We tested whether these entities are mechanically non-equivalent in vivo and separable by MRE-derived viscoelastic metrics and perilesional interface-instability features.

**Methods:** In a prospective, histopathology-anchored cohort, 11 post-radiotherapy enhancing lesions were classified as RN (n=3) or recurrent/progressive tumor (n=8). MRE was acquired at 3.0 T with single-frequency 60-Hz excitation to derive storage modulus (G′), loss modulus (G″), and complex shear modulus magnitude (|G*|). Co-primary endpoints were median tumor G′ and |G*|, each tested one-sided (RN > tumor) with Holm correction across the two co-primary tests. Median tumor G″ was tested two-sided. A prespecified secondary 6-endpoint family (absolute and tumor/NAWM-normalized G′, G″, and |G*|) was analyzed with Benjamini–Hochberg FDR control. Exploratory instability mapping in a 0–6 mm peritumoral shell generated interface-topology metrics, including convexity.

**Results:** Absolute tumor-core medians were higher in RN than tumor for |G*| (1.79 vs 1.32 kPa; Cliff’s δ=0.67; q=0.10), G′ (1.62 vs 1.09 kPa; δ=0.50; q=0.14), and G″ (0.81 vs 0.46 kPa; δ=0.75; q=0.10). NAWM normalization improved separation: tumor/NAWM |G*| (2.26 vs 1.41; δ=0.92; q=0.04) and tumor/NAWM G″ (2.67 vs 0.87; δ=1.00; q=0.04) were FDR-significant. Convexity also differentiated RN from tumor (0.49 vs 0.36; δ=1.00; MWU p=0.01).

**Conclusions:** Tumor/NAWM G″, tumor/NAWM |G*|, convexity, and tumor G″ emerged as the strongest candidate features, indicating that RN is mechanically harder and more dissipative than recurrent metastasis. Signal strength was high (Cliff’s δ up to 1.00) but should be interpreted cautiously given sample size. Exploratory analyses further suggest that instability mapping captures biologically relevant interface behavior. These findings support a mechanics-based RN-versus-recurrence framework and justify prespecified, preregistered external validation.

## Introduction

Post-radiotherapy enhancing lesions in patients with brain metastases remain a high-impact diagnostic dilemma because conventional post-contrast MRI often cannot reliably distinguish radiation necrosis (RN) from true tumor progression (TTP)^1–3^. RN and TTP frequently present with overlapping appearances on conventional MRI, yet imply different management pathways. Recurrence commonly triggers treatment escalation and/or surgical re-intervention, whereas RN is often approached with surveillance, corticosteroids, and in selected cases anti-VEGF therapy^1,4,5^. Although advanced imaging and multidisciplinary interpretation improve decision-making, diagnostic uncertainty persists in a clinically meaningful subset of cases, often necessitating serial follow-up and, in many patients, invasive tissue confirmation^6–9^. The ability to reliably distinguish these entities non-invasively would have clear clinical implications for treatment selection, procedural burden, and risk exposure.

The diagnostic ambiguity arises because structurally similar post-treatment enhancement can reflect biologically distinct states. Radiation necrosis is a delayed radiation-injury syndrome characterized by vascular injury, blood–brain barrier disruption, neuroinflammation, and matrix remodeling, whereas recurrence represents viable neoplasm with dynamic tumor–stroma coupling and invasive interface behavior^10–12^. These entities may converge radiologically on conventional MRI, but there is no biological reason to assume that they are mechanically equivalent. Contrary, in operative practice, lesions later confirmed as RN are often perceived as harder and more fibrotic than recurrent tumours. This premise is consistent with mechanobiological studies showing that extracellular matrix stiffness and viscoelasticity influence cell-state programs, migration phenotypes, and tissue architecture across disease systems^13–19^. By the same logic, interface-physics frameworks predict that lesions with infiltrative, heterogeneously coupled boundaries should generate different spatial viscoelastic organization than lesions dominated by treatment-related consolidation^10,20–22^. Together, these considerations support a mechanics-first hypothesis: RN and recurrent metastasis may be better separated by viscoelastic state and perilesional interface organization than by morphology alone.

Magnetic resonance elastography (MRE) provides a direct in vivo readout of tissue mechanics via the complex shear modulus, including storage (G′), loss (G″), and magnitude (|G*|) components^23,24^. In post-radiotherapy lesions, this decomposition is particularly relevant because fibrosis, necrosis, edema, inflammation, and residual tumour are expected to alter elastic energy storage and viscous dissipation differentially. Interface stability refers to the mechanical coherence of the lesion–brain boundary under physiologic loading. When adjacent regions have strong viscoelastic contrast, resulting stress/strain concentration at the interface may create microenvironmental cues that facilitate invasive behavior, and should manifest in MRE as spatially structured perilesional heterogeneity rather than only changes in mean or median values^10^.

Accordingly, this histopathology-anchored lesion-level study was designed around three prespecified objectives. First, it tested whether RN exhibits higher tumour viscoelastic magnitude than recurrence for |G*| and G′, using directional one-sided inference (RN > recurrence), while evaluating G″ with a two-sided test because no directional prior was imposed for dissipation. Second, it assessed whether normalization to contralateral normal-appearing white matter (NAWM) improves class separation by attenuating between-scan baseline variability. Third, it examined instability-oriented perilesional spatial mapping as an exploratory interface readout to determine whether relative spatial organization contributes information beyond regional summaries.

## Methods

### Study design and cohort

This prospective study was approved by the national ethical committee (ID: 2400648) and performed according to the Declaration of Helsinki. Adult patients with radiotherapy-treated cerebral metastases and suspected TTP on imaging were offered participation. All patients underwent preoperative MRI and MRE. Lesions were categorized as RN or recurrence on histopathology according to current criteria (Figure 1).

**Figure 1.**
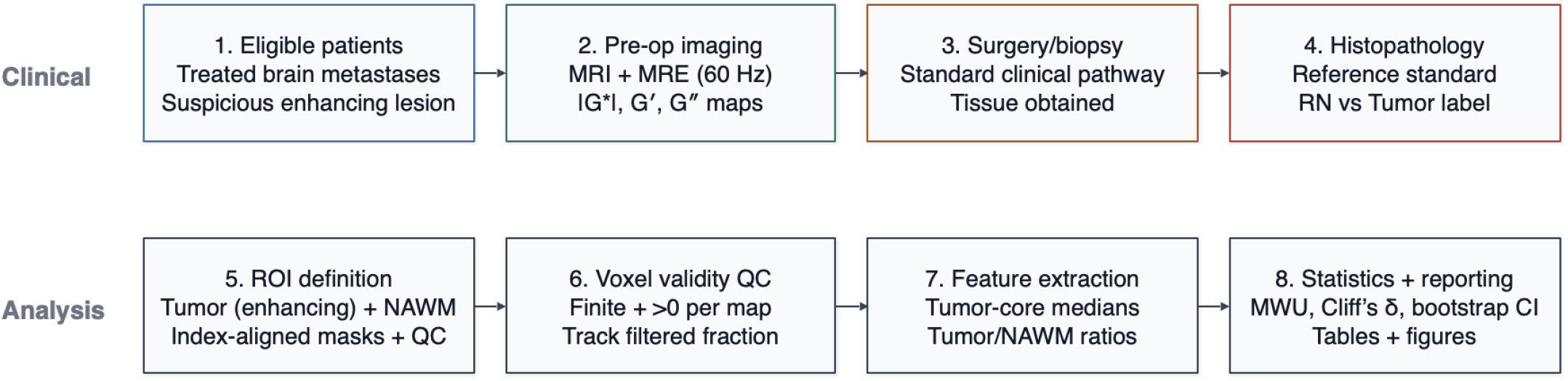
Flowchart

### Imaging data and derived MRE parameter maps

MRE was acquired on a 3.0-T scanner (Philips Achieva, Philips Healthcare, the Netherlands) with a 16-channel head coil^25^. The protocol also included post-contrast T1-weighted gradient-echo imaging after intravenous gadobutrol (Gadovist, 1 mmol/mL; Bayer, Germany) at a dose of 0.1 mL/kg. Mechanical excitation was applied at 60 Hz using a pneumatic actuator system (Resoundant Inc., USA) placed under the head, with drive amplitude set to approximately 20% of maximum output^25^. Wave-motion data were collected with a single-shot spin-echo EPI MRE sequence (TR/TE 4800/67 ms; FOV 240 × 240 mm^2^; SENSE factor 3; 48 slices; 3-mm slice thickness; isotropic 3-mm voxels; 8 phase offsets).

Elastographic post-processing used direct inversion to generate parametric maps of storage modulus (G′), loss modulus (G″), and complex shear modulus magnitude (|G*|)^25^. All parameter maps were rigidly aligned and resampled onto the G′ reference grid in 3D Slicer (v5.7), followed by visual quality control.

### Segmentation and anatomical regions

Lesion and tissue masks were created in 3D Slicer. The tumour ROI was defined by contrast-enhancing lesional voxels on post-contrast T1-weighted MRI. Peritumoral tissue was defined as the immediate non-enhancing tissue compartment surrounding the enhancing lesion according to the prespecified segmentation workflow used for ROI extraction. NAWM was defined as a contralateral spherical ROI (6 mm diameter) placed in the hand-knob white matter region, avoiding visible pathology. Labelmaps were exported as NIfTI volumes intended to match the MRE grid and were used for all ROI-based feature extraction. Visual verification of ROI alignment was performed on QC panels.

All quantitative analyses were performed on one reference grid per lesion, using the G′ grid as canonical reference when combining G′, G″, |G*|, segmentations, and instability maps. MRE maps may contain non-finite values and, particularly for G″, non-physical values (≤0) due to inversion noise/reconstruction artifacts. Voxels were considered valid only if finite and strictly positive for the parameter analyzed (|G*|, G′, and G″: finite and >0).

### ROI-based viscoelastic feature extraction

For each lesion, region (tumor, NAWM), and modality (|G*|, G′, G″), distribution summaries were computed using valid voxels only; lesion size was quantified from the tumor mask and converted to volume (mm^3^) using the analysis voxel volume. Summaries included mean, median, SD, IQR, selected percentiles, minimum, and maximum.

In addition to absolute values, NAWM-normalized tumor ratios were computed as:

*Tumour* / *NAWM* =*median*(*tumour*)/ *median*(*NAWM*),

### Instability mapping in a peritumoral annulus

To characterize spatially organized viscoelastic imbalance at the tumor–brain interface, an instability index I(x) was evaluated within a three-dimensional peritumoral shell extending 0–6 mm from the tumor boundary^10^. A separate 3-mm peritumoral rim directly adjacent to the tumor was defined as the reference region.

Tumor masks were first hole-filled in 3D, after which Euclidean distance-to-boundary maps (in millimeters) were computed using voxel spacing from the G′ image header. The analysis shell comprised voxels outside the tumor with boundary distance in the interval (0, 6] mm. The reference rim comprised voxels in (0, 3] mm from the boundary, corresponding to approximately one voxel at the native 3-mm resolution. In practice, the rim definition was implemented either as a one-voxel dilation band or with an equivalent distance-threshold formulation, yielding the same geometric target region.

Let tan (*δ*)(*x*)=*G″* (*x*)/*G ′* (*x*). With reference medians G″r,med and tan(δ)r,med measured in the rim, instability was defined as:

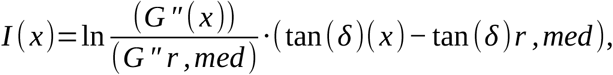

computed where *G′* (*x*)>0, *G″* (*x*)>0, and tan (*δ*)(*x*) is finite^10^.

From shell I(x), the analysis computed central/tail summaries (median, p95), threshold fractions (I>0.02, I>0.05, I<0), Shannon entropy (32 bins over fixed range [0, 0.4]), positive-tail AUC (capped at Imax=0.30), and radial profile/radial AUC (0–2, 2–4, 4–6 mm).

With unstable mask *I* (*x*)>0.02, per-slice topology was computed (when unstable area >0): isoperimetric ratio (*IPR* : *P*^2^ / 4 *πA*), convexity (*A* / *Ahull*), skeleton length density (*nskel* / *A*), and skeleton branch density (*nbranch* / *nskel*). Slice metrics were area-weighted across slices.

### Statistical analysis

Because of small sample size and non-Gaussian distributions, group comparisons used nonparametric inference. For each metric, the analysis reports Mann–Whitney U p-values, Cliff’s delta (RN minus Tumor direction), and bootstrap 95% CIs (20,000 resamples) for median difference (RN − Tumour).

Primary inference used directional tumor co-primary endpoints (absolute G′ and absolute |G*|), each tested one-sided for RN>Tumor and adjusted with Holm correction across the two co-primary tests. G″ was analyzed with two-sided inference because no directional prior was imposed.

Secondary tumor endpoint family included six metrics: absolute G′, absolute G″, absolute |G*|, tumor/NAWM G′, tumor/NAWM G″, and tumor/NAWM |G*|. False-discovery control for this family used Benjamini–Hochberg FDR. Exploratory instability analyses were interpreted as hypothesis-generating using effect sizes and uncertainty intervals rather than as confirmatory endpoints. Given the expected dependence among these metrics, FDR control is used to limit false discoveries while retaining power in this exploratory secondary family.

Standard Cliff’s delta thresholds were used: negligible <0.147, small 0.147–<0.33, medium 0.33–<0.474, large ≥0.474^26^.

### Software and robustness

All analyses were performed in Python (NumPy, Pandas, NiBabel, SciPy, scikit-image, Matplotlib) in a scripted notebook workflow. Segmentations were produced in 3D Slicer. Intermediate masks, instability maps, QC panels, and summary tables were exported to structured directories for full traceability. We performed a robustness analysis of the instability pipeline by varying two parameters: the peritumoral shell definition (outer radius 4, 6, and 8 mm from the tumor boundary) and the instability binarization threshold (τ = 0.015, 0.020, 0.030, and 0.050). For each parameter combination, we recomputed the same interface-derived metrics and summarized between-group separation (RN vs recurrent/progressive tumor) using median differences (RN − tumor) and Cliff’s delta. This analysis was designed to test whether observed group contrasts were stable to plausible choices of rim/shell geometry and instability cutoff.

## Results

A total of 11 post-radiotherapy enhancing lesions were included (RN n=3; recurrent/progressive tumor n=8) (Table 1). Primary analyses were performed at lesion level across |G*|, G′, and G″, using tumor absolute values and NAWM-normalized contrasts. Mean lesion size was 506 mm^3^ (range 171–1161) in radiation necrosis and 1297 mm^3^ (range 396–3831) in the tumor/recurrent group (Table 2).

**Table 1:**
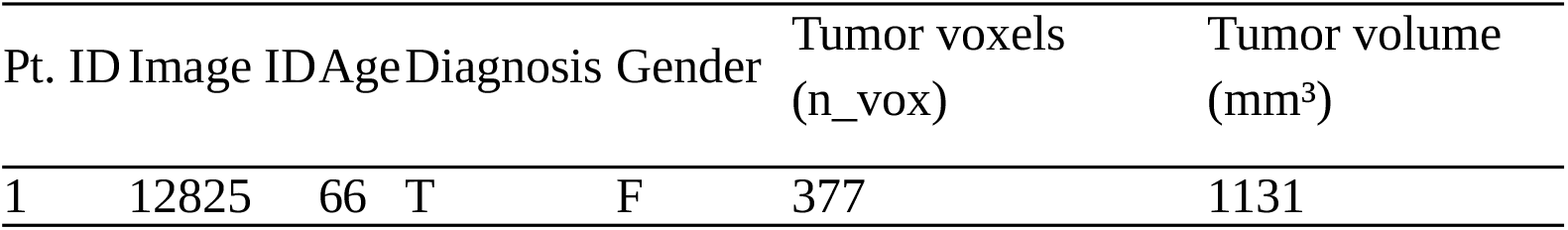
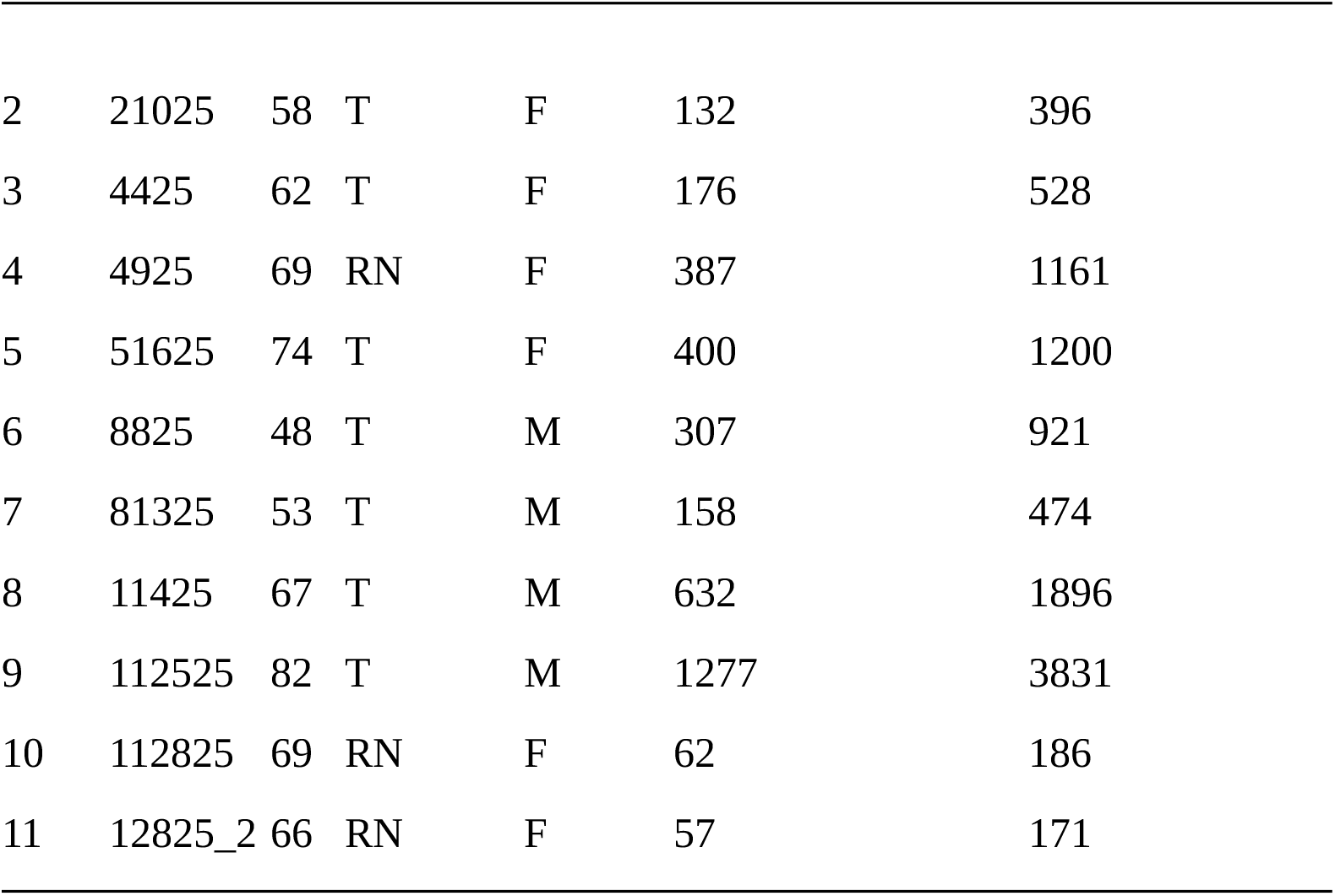
Cohort charateritics.

**Table 2:**
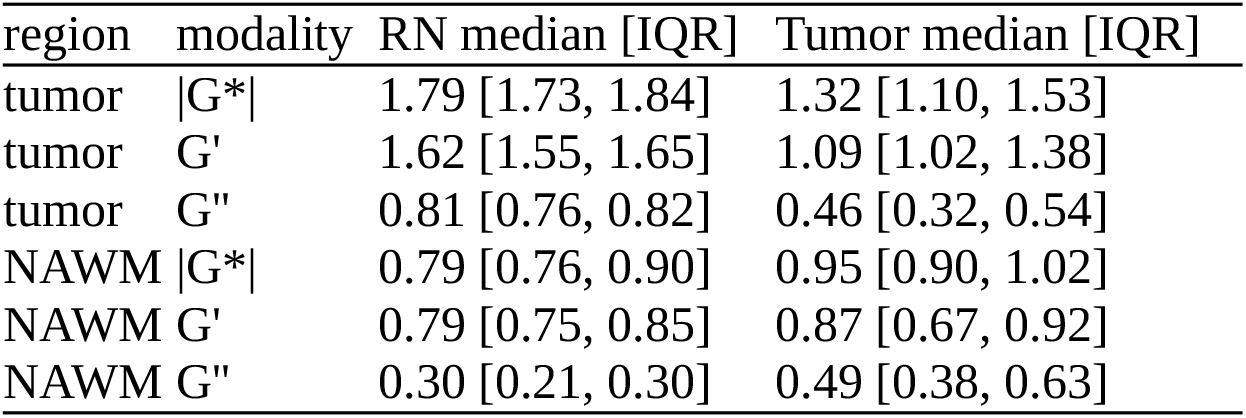
Summary statistics.

Descriptive viscoelastic summaries showed a consistent directional pattern (Table 2). In tumor core, RN had higher medians than tumor for all three parameters: |G*| (1.79 vs 1.32 kPa), G′ (1.62 vs 1.09 kPa), and G″ (0.81 vs 0.46 kPa). In contralateral NAWM, medians trended lower in RN than in tumor (| G*|: 0.79 vs 0.95; G′: 0.79 vs 0.87; G″: 0.30 vs 0.49), which amplified lesion-to-background contrast in normalized analyses.

Formal tumor between-group testing of absolute parameters (Table 3, Figure 2) was directionally concordant with expectations and showed large effect sizes by Cliff’s δ for all modalities: |G*| δ=0.67 (large), G′ δ=0.50 (large), and G″ δ=0.75 (large). Median differences were 0.48 kPa for |G*|, 0.54 kPa for G′, and 0.35 kPa for G″ (RN−Tumor). After multiplicity correction within the absolute 3-endpoint block, none reached adjusted significance (q=0.10, q=0.14, and q=0.10, respectively).

**Table 3:**
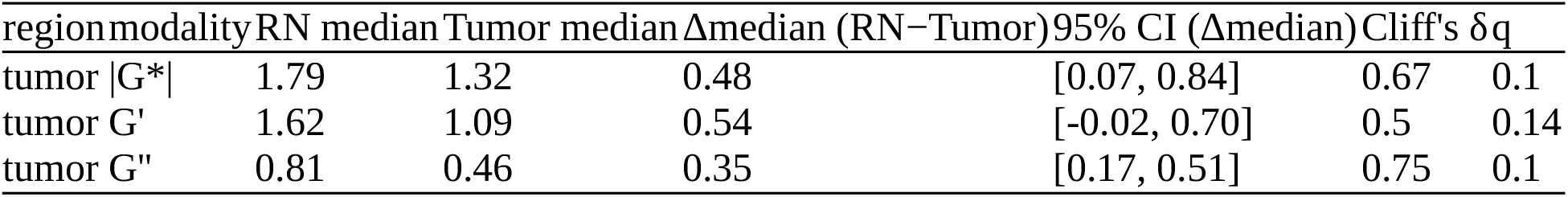
Statistical summary raw viscoelastic parameters by groups.

**Figure 2:**
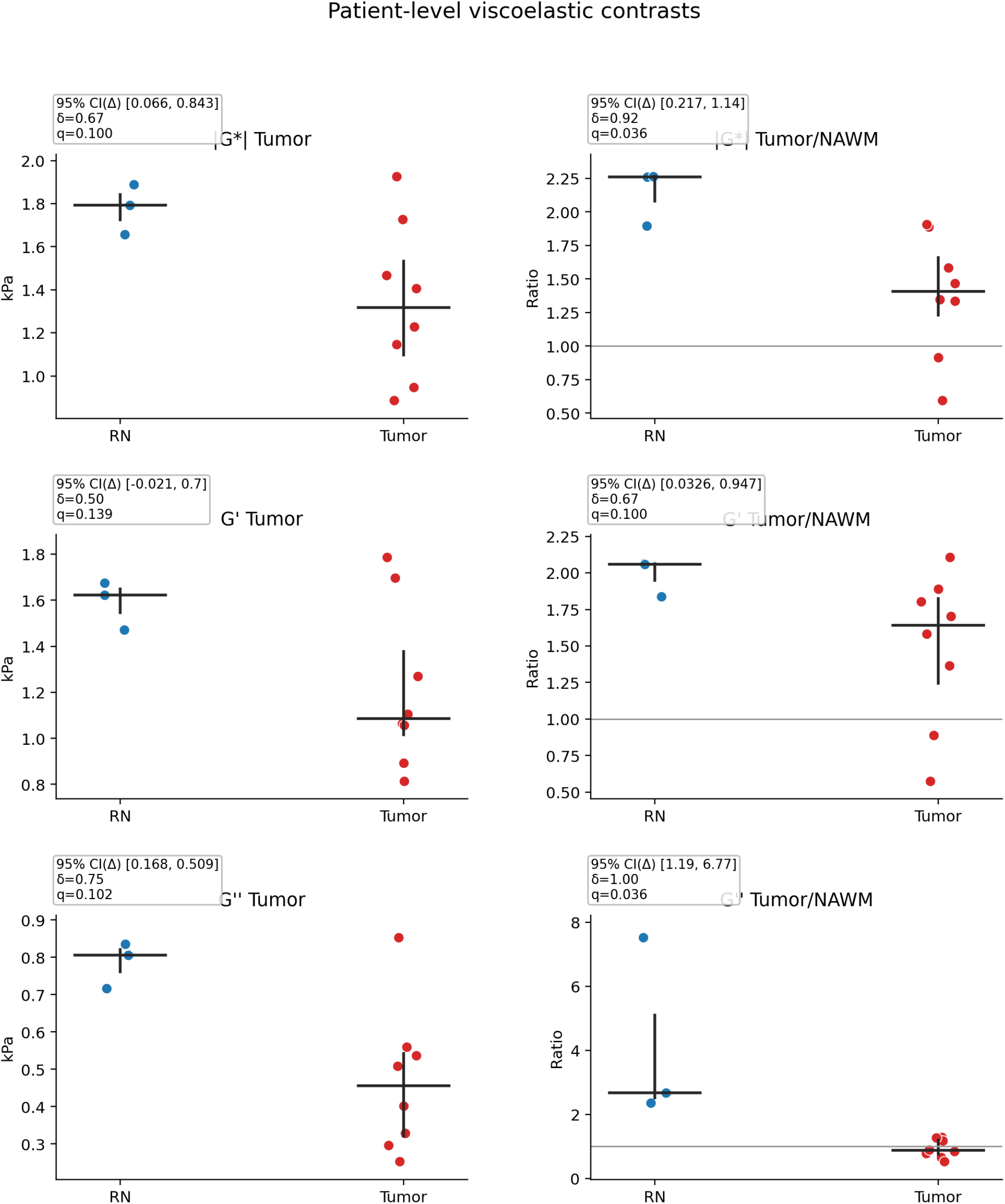
Viscoelastic contrast by group

NAWM-normalized analyses (Table 4, Figure 2) retained the same directionality and increased separation. Effect sizes were again large across all normalized endpoints: tumor/NAWM |G*| δ=0.92 (large), tumor/NAWM G′ δ=0.67 (large), and tumor/NAWM G″ δ=1.00 (large; complete rank separation). Under all-6 FDR control, normalized |G*| and normalized G″ were statistically significant (both q=0.04), whereas normalized G′ did not meet threshold (q=0.10). Median normalized contrasts were 2.26 vs 1.41 for |G*|, 2.06 vs 1.64 for G′, and 2.67 vs 0.87 for G″ (RN vs tumor), with RN−Tumor median differences of 0.86, 0.42, and 1.80, respectively.

**Table 4:**
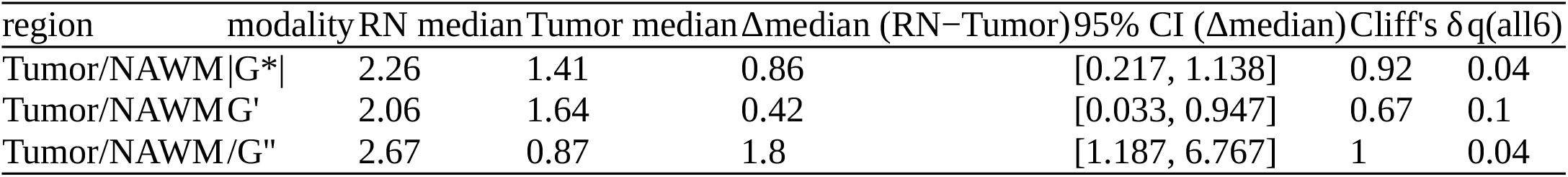
Statistical summary NAWM normalized viscoelastic parameters by group.

Exploratory instability metrics (Table 5; Figure 3) showed nonuniform behavior across features. Convexity demonstrated the strongest and clearest separation, with a large effect size (δ=1.00, large) and two-sided MWU p=0.01. Isoperimetric ratio also showed a large effect (δ=−0.67, large), and skeleton branch-point density showed a large positive effect (δ=0.75, large), but these did not achieve statistical significance in this sample. Tail-AUC and skeleton length density showed small effects (δ=0.17 each), while radial-AUC and entropy were negligible to small (δ=0.00 and δ=−0.17, respectively). As prespecified, this instability block is interpreted as hypothesis-generating.

**Table 5:**
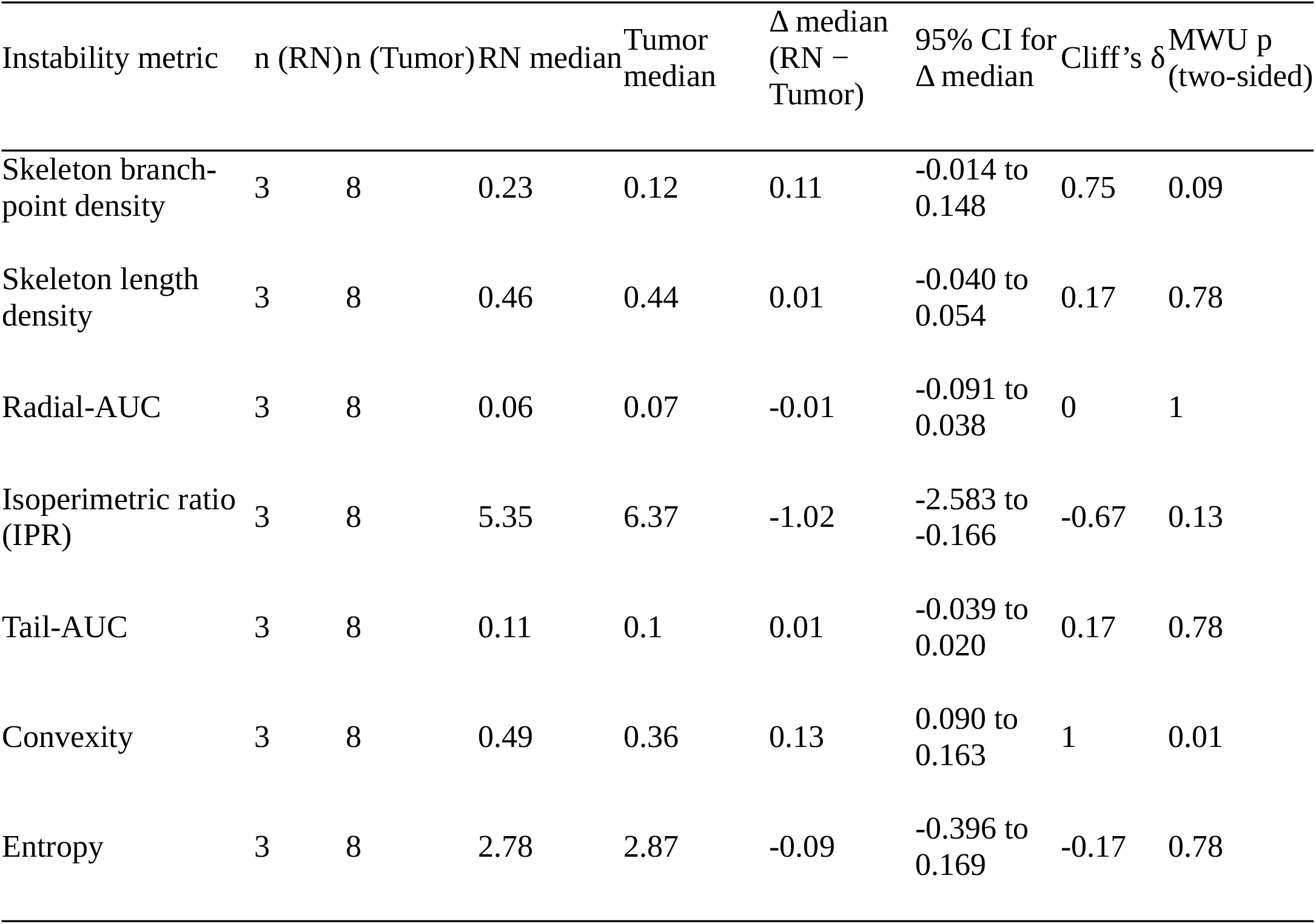
Summary of instability metrics.

**Figure 3:**
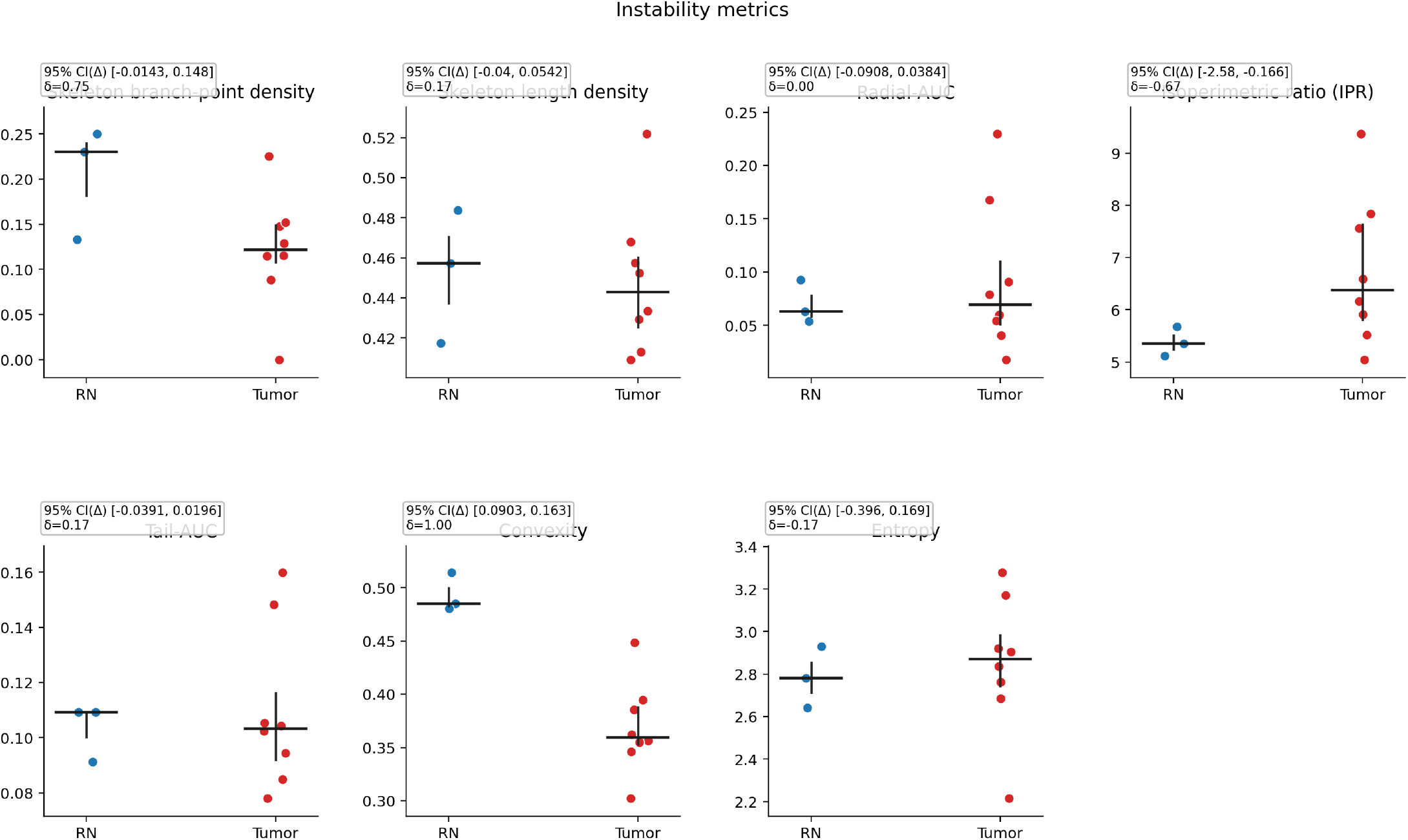
Instability metrics by group

Taken together, the results indicate a coherent and directionally stable biomechanical signal separating RN from recurrent/progressive tumor (Figure 4). The primary absolute tumor-core endpoints showed uniformly large effect sizes but did not survive multiplicity adjustment, whereas the strongest inferential support came from NAWM-normalized |G*| and G″, both with large effects and FDR-significant q-values. Exploratory instability readouts added complementary spatial information, with convexity as the most prominent candidate for follow-up validation.

**Figure 4:**
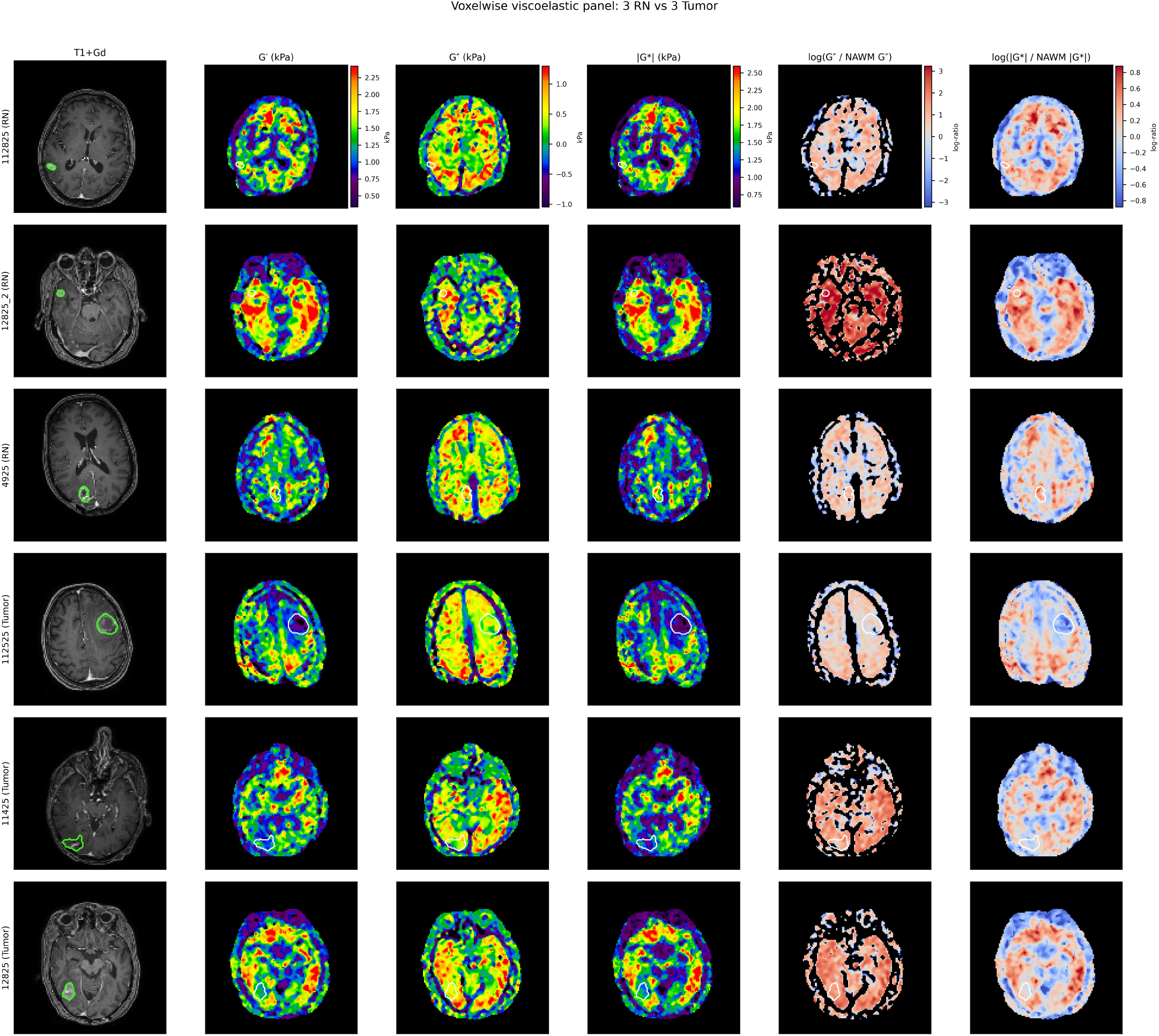
Voxelwise viscoelastic maps in post-radiotherapy enhancing lesions with histopathology-confirmed diagnosis. Representative cases from the analysis cohort are shown (3 radiation necrosis (RN), top rows; 3 recurrent/progressive tumor, bottom rows). Columns display post-contrast T1-weighted MRI (T1+Gd), storage modulus G′ (kPa), loss modulus G″ (kPa), complex shear modulus magnitude |G*| (kPa), and NAWM-normalized log-ratio maps for dissipation and stiffness (log(G″/NAWM G″), log(|G*|/NAWM |G*|)). Tumor contours are overlaid (green on T1+Gd; white on viscoelastic maps). MRE parameter maps (G′, G″, |G*|) are rendered with a common\ colormap and column-wise pooled robust intensity limits to preserve direct cross-patient comparability within each parameter. Normalized maps are rendered with a diverging colormap centered at zero (log-ratio), where warm colors indicate values above and cool colors values below NAWM reference. Background is masked to black, and MRE units are harmonized to kPa before visualization. This panel is intended as a qualitative spatial illustration of lesion-level mechanical phenotype; inferential statistics are reported separately in Tables 3–4.

### Sensitivity and robustness

Across the tested range of τ values, effect estimates were unchanged within each shell definition for the reported metrics, indicating threshold robustness in this cohort. Shell-definition effects were metric dependent: annulus voxel burden remained directionally consistent across all shell radii (RN lower than tumor; Cliff’s δ approximately −0.58 to −0.67), whereas convexity showed greater dependence on shell size, with near-null separation at 4 mm (δ ≈ −0.08) and clearer RN-dominant separation at 6 mm (δ ≈ +0.42). Overall, these findings support robustness of size-related interface measures and suggest that geometric topology descriptors are sensitive to shell parameterization and may require standardized shell definitions in validation cohorts.

## Discussion

This histopathology-anchored pilot demonstrates a coherent biomechanical separation between RN and recurrent/progressive metastasis at lesion level, with the clearest signal emerging in tumor-core viscoelasticity and lesion-to-background normalization. Across absolute tumor-core measurements, RN showed higher central values for all three viscoelastic parameters, and NAWM normalization strengthened separation for the most informative endpoints, particularly those reflecting dissipative behavior. The increased signal form normalization is in contrast with our earlier study, however, in this cohorts NAWM was defined in a defined anatomical location instead of mirroring the lesion^27^.

A biologically coherent hierarchy is visible. That ordering is mechanistically credible for delayed radiation injury, where extracellular-matrix remodeling, microvascular dysfunction, and chronic inflammatory restructuring are expected to perturb viscous energy dissipation alongside elastic storage. The immediate observable effect is that RN lesions appear harder than metastases during surgery. From an inferential standpoint, the staged endpoint logic remains important. The directional co-primary framework was therefore biologically consistent, but the study should still be interpreted conservatively at this sample size. The most robust statistical signal in the current dataset sits in the normalized family, not as a standalone claim of finalized diagnostic performance. This distinction is methodologically important: the present results support prioritization and design of validation endpoints, rather than immediate clinical deployment thresholds.

The exploratory instability domain is mechanically plausible as a readout of boundary organization, where instability is a feature of invasive phenotypes^10^. In interface-physics terms, both Hele–Shaw– type fingering analogies and Biot-style surface-instability concepts imply that boundary geometry is governed by local contrasts in coupling, dissipation, and stress transfer across adjacent media^20,22,28^. The framework explains why perilesional contrast may discriminate RN from recurrent tumor based on interphase dynamics. In this dataset, convexity provided the clearest RN-dominant separation. This suggests more irregular interface geometry around metastases. IPR and branch-density shifted in different directions, consistent with these metrics capturing different geometric attributes of the interface (global compactness versus branching complexity). This pattern is also in line with results from our earlier instability study, where boundary-organization features carried signal beyond central tendency summaries, and instability increased with WHO grade^10^. Other instability features were smaller or less precise, which is expected in a low-n topology setting. These descriptors are therefore best interpreted as mechanistically grounded, hypothesis-generating candidates that should be carried forward into prespecified validation.

Compared with conventional MRI-based RN-versus-recurrence markers, the rank-separation magnitudes observed here are encouraging^6,8,9^. In the current cohort, normalized viscoelastic endpoints reached very large nonparametric effects (δ 0.92 for tumor/NAWM |G*|; δ 1.00 for tumor/NAWM G″). That does not imply that MRE has solved the diagnostic problem, and it should not be interpreted as direct superiority without same-cohort head-to-head modeling against perfusion, spectroscopy, and advanced structural/radiomic pipelines. It does, however, indicate that the biomechanical signal strength is competitive and, in this dataset, potentially stronger than many single-feature legacy approaches typically used in isolation.

Clinically, the immediate implication is prioritization rather than deployment. A rational next step is a pre-registered, adequately powered validation study with locked preprocessing/QC, fixed endpoint hierarchy, and predefined decision thresholds. The most defensible candidate structure from the present data is a multi-domain model combining (1) absolute tumor-core viscoelastic state, (2) normalized tumor-core contrast with emphasis on G″ and |G*|, and (3) exploratory instability topology as an adjunctive spatial layer. Prospective same-patient benchmarking against contemporary MRI biomarker stacks is necessary before translational claims about routine decision support.

The limitations are clear and should remain explicit. The RN group is small (n=3), which limits precision and can inflate apparent effect-size volatility despite the directional consistency being strong. Confidence intervals are narrow for some high-signal metrics (for example convexity and normalized G″), but many exploratory metrics retain broad uncertainty and several cross zero. ROI definition and instability-threshold dependencies can influence topology outputs. In addition, NAWM normalization may reduce unwanted inter-scan variance while also absorbing biologically relevant intersubject information, which will need targeted analysis in larger cohorts.

## Conclusion

In this histopathology-anchored pilot, RN and recurrent/progressive metastasis showed strong and consistent biomechanical differences on MRE, with the most robust separation in NAWM-normalized parameters. While absolute tumor-core metrics were directionally concordant (higher RN medians for | G*|, G′, and G″), the strongest inferential signal was observed for normalized tumor/NAWM |G*| and G″ (both q=0.04), with large to maximal effect sizes (Cliff’s δ=0.92 and 1.00, respectively). Among exploratory interface-topology features, convexity provided the clearest RN-dominant separation (δ=1.00; p=0.01), whereas other topology descriptors were weaker or less precise, consistent with small-sample exploratory behavior.

Taken together, these findings support a mechanics-based framework in which dissipation-sensitive and normalized viscoelastic contrasts are the most promising candidate biomarkers for RN-versus-recurrence discrimination. At the same time, the small RN subgroup and exploratory nature of topology analyses require cautious interpretation. The immediate implication is prioritization of locked, pre-registered external validation—centered on normalized |G*| and G″, with convexity as an adjunct spatial marker—rather than direct clinical deployment at this stage.

## Data Availability

All data produced in the present study are available upon reasonable request to the authors

## Acknowledgements

We acknowledge Dr. Sandeep Ganji at Philips Healthcare and Dr. Yuan Le at Mayo Clinic for developing the imaging pulse sequence, establishing imaging protocols and processing some of the images for this study. The study was supported by the Danish Cancer Society and NIH.

